# Paratyphoid Fever and the Genomics of *Salmonella enterica* Serovar Paratyphi A in Taiwan

**DOI:** 10.1101/2025.04.11.25325632

**Authors:** Ying-Shu Liao, Yu-Ping Hong, Bo-Han Chen, You-Wun Wan, Ru-Hsiou Teng, Shiu-Yun Liang, Hsiao Lun Wei, Jui-Hsien Chang, Ming-Hao Yang, Chi-Sen Tsao, Chien-Shun Chiou

## Abstract

**Background:** *Salmonella enterica* serovar Paratyphi A has emerged as a significant global health concern due to the progressive development of antimicrobial resistance and its broader geographic distribution. In Taiwan, paratyphoid fever was historically rare and predominantly associated with imported cases. Since 2022, however, a marked increase in domestically acquired infections has been observed, prompting investigations into their origin and likely route of introduction.

**Methods:** We analyzed surveillance data on 223 patients with paratyphoid fever reported in Taiwan between 2001 and 2024. Whole-genome sequencing and antimicrobial susceptibility testing were performed on 88 *S.* Paratyphi A isolates obtained from both imported and domestically acquired infections from 2007 to 2024. Phylogenetic analysis and genotyping were conducted to assess genetic relatedness and to trace potential sources of introduction by comparing them with global isolates.

**Results:** Although 55.2% of paratyphoid fever infections were imported, domestically acquired infections became predominant after 2022. Most isolates (76.1%) were resistant to nalidixic acid and nonsusceptible to ciprofloxacin due to *gyrA* mutations at codon 83 (S83F or S83Y). The majority of domestic isolates were classified as ST129 and paratype 2.4 and showed close genetic relatedness to strains from Indonesia. Of the 31 domestic isolates collected between 2022 and 2024, 30 clustered with Indonesian strains, and 28 exhibited very limited genetic divergence, suggesting a likely common source of infection.

**Conclusions:** *S. Paratyphi A* isolates recovered from recent domestically acquired infections in Taiwan are genetically related to strains from Indonesia. The high rate of ciprofloxacin nonsusceptibility, driven by *gyrA* mutations, highlights the need for ongoing resistance monitoring, while the preserved susceptibility to traditional first-line agents supports their continued use in treatment. Genomic surveillance proved effective for source tracking and guiding public health interventions.

**Author summary:** Paratyphoid fever, caused by *Salmonella enterica* serovar Paratyphi A, is typically associated with endemic regions in South and Southeast Asia. In Taiwan, the disease has historically been rare and primarily linked to imported cases. However, since 2022, the number of locally acquired infections has increased. We analyzed 88 *S.* Paratyphi A isolates collected in Taiwan between 2007 and 2024 using whole-genome sequencing and antimicrobial susceptibility testing. While 76.1% of isolates were resistant to nalidixic acid and exhibited reduced susceptibility to ciprofloxacin, these phenotypes were associated with gyrA mutations at codon 83, which are known to impair the efficacy of fluoroquinolones. Importantly, all isolates remained susceptible to 13 other tested antimicrobials, including older drugs such as ampicillin, chloramphenicol, and trimethoprim-sulfamethoxazole. Phylogenetic analysis showed that most domestic isolates belonged to the same genetic lineage as strains from Indonesia. Notably, 28 of 30 isolates collected from domestic cases between 2022 and 2024 exhibited near-identical genetic profiles, despite patients having no epidemiological links. This suggests that the infections likely originated from a common external source, such as contaminated imported food or an environmental reservoir, rather than through sustained local transmission. Our study highlights the value of genomic surveillance in tracing infection sources and provides evidence to support targeted public health interventions and treatment strategies that preserve the use of traditional first-line antibiotics where susceptibility remains high.

## Introduction

Paratyphoid fever is a systemic illness caused by *Salmonella enterica* serovars Paratyphi A, B, and C, with *S.* Paratyphi A being the predominant cause globally. In 2017, an estimated 3.4 million cases were reported worldwide [1]. Although the overall case fatality rate is under 1% with proper treatment, complications such as intestinal perforation and septicemia can increase mortality, especially in areas with limited healthcare access. The disease, primarily transmitted through the ingestion of contaminated food or water, exhibits a high prevalence in regions with inadequate sanitation, including South and Southeast Asia, sub-Saharan Africa, and Oceania. Within these endemic regions, countries such as India, Nepal, Pakistan, and parts of China have seen *S.* Paratyphi A become a notable cause of enteric fever, sometimes exceeding the prevalence of *S.* Typhi [2]. In contrast, paratyphoid fever cases reported in high-income countries, including Australia, Japan, and European countries, are predominantly travel-related, with most infections traced to travelers returning from endemic areas [2–4].

The increasing prevalence of antimicrobial-resistant *S.* Paratyphi A strains, especially those with reduced susceptibility to fluoroquinolones, presents a significant clinical concern [5]. While most isolates remain susceptible to first-line agents such as ampicillin, chloramphenicol, and cotrimoxazole, as well as third-generation cephalosporins and macrolides, nonsusceptibility to fluoroquinolones is commonly observed and often linked to mutations in the *gyrA* gene, particularly at codon 83 (S83F and S83Y) [6–8]. The widespread presence of these mutations across diverse geographic settings highlights the importance of resistance monitoring through genomic surveillance.

Recent phylogenomic analyses, including whole-genome sequencing of outbreak isolates from India, Bangladesh, and China, have significantly enhanced our understanding of *S.* Paratyphi A transmission dynamics and population structure [6, 8, 9]. These studies have revealed localized outbreaks and suggested cross-border circulation of predominant genotypes, thereby contributing to the rising disease burden in South and Southeast Asia. To facilitate comparative genomic analysis, Tanmoy et al. developed Paratype, an SNP-based genotyping tool that enables standardized classification of *S.* Paratyphi A clades and genotypes [10]. Using this system, studies in Bangladesh, India, Cambodia, and China have identified the predominance of globally distributed lineages demonstrating intranational genetic homogeneity but marked interregional diversity [6–9].

In Taiwan, paratyphoid fever is a notifiable disease with historically low incidence, primarily imported from endemic regions [11]. However, a notable increase in locally acquired infections since 2022 has raised concerns about ongoing domestic transmission. To clarify the origin and transmission dynamics of these recent cases, we analyzed the genomic and antimicrobial resistance profiles of *S.* Paratyphi A isolates from both imported and domestic infections and compared them with international strains.

## Methods

### Epidemiological data of paratyphoid fever

Paratyphoid fever is a notifiable disease in Taiwan, hospitals are required to report cases to health authorities and submit isolates to the Taiwan Centers for Disease Control (Taiwan CDC). Isolates were confirmed as *Salmonella* using the Bruker MALDI Biotyper, and their serotypes were determined according to the method developed by Chiou et al. [12]. Statistical data on reported paratyphoid cases from 2001 to 2024 were obtained from the Taiwan National Infectious Disease Statistics System (NIDSS; https://nidss.cdc.gov.tw/en/Home/Index). Demographic information, including sex, age, country of citizenship, country of residence, travel history, and year of onset, was retrieved from the Taiwan National Notifiable Disease Surveillance System with authorization from the Taiwan CDC (IRB113107#1).

### Antimicrobial susceptibility testing (AST)

Isolates were tested for antimicrobial susceptibility with the EUVSEC3 Sensititre MIC panel (TREK Diagnostic Systems Ltd., West Essex, England), following the manufacturer’s instruction. The MIC breakpoints for *Enterobacterales*, as defined by the Clinical and Laboratory Standards Institute 33rd edition [13], were used to interpret the antimicrobial susceptibility testing results for amikacin, ampicillin, azithromycin, cefotaxime, ceftazidime, chloramphenicol, ciprofloxacin, colistin, gentamicin, meropenem, nalidixic acid, sulfamethoxazole, tetracycline, and trimethoprim. The MIC results for tigecycline were interpreted using the standard set by the European Committee on Antimicrobial Susceptibility Testing (EUCAST) [14].

### Whole genome sequencing (WGS)

We obtained 88 *S.* Paratyphi A isolates recovered between 2007 and 2024 for WGS to identify resistance determinants and analyze genomic characteristics. WGS was conducted using the Illumina MiSeq platform (Illumina Inc., California, USA). Illumina sequence reads were assembled using SPAdes version 3.15.3 [15]. The assembled sequences (contigs) were analyzed using the AMRFinderPlus pipeline provided in the NCBI database [16], ResFinder and PlasmidFinder supplied by the Center for Genomic Epidemiology (http://www.genomicepidemiology.org), and Paratype software tool (https://github.com/CHRF-Genomics/Paratype/) [10], to determine antimicrobial resistance determinants, plasmid incompatibility (Inc) types, paratypes, and conventional sequence types (STs).

Phylogenetic relationships among *S.* Paratyphi A isolates were inferred using core genome single nucleotide polymorphism (cgSNP) profiles. SNP calling was performed by generating a multiple-sequence alignment file, where contigs of each isolate were mapped against the reference genome *S.* Paratyphi A AKU12601 (GenBank accession: FM200053) using ska.rust v0.3.7 (https://github.com/bacpop/ska.rust) [17]. To eliminate confounding factors in phylogenetic reconstruction, repetitive regions, prophages, and recombinant regions were excluded using BEDTools (https://github.com/arq5x/bedtools2) [18] with the PARAREPEAT file (https://github.com/katholt/typhoid). The filtered alignment was then used for cgSNP profile calling with Gubbins v3.3.1 (https://github.com/nickjcroucher/gubbins) [19]. Finally, the phylogenetic tree was visualized and annotated using iTOL (https://itol.embl.de) [20].

### Global comparison of *S.* Paratyphi A isolates

Isolates from Taiwan were compared with those from Bangladesh [6, 21], Cambodia [7, 22], China [9], India [8, 23, 24], Indonesia [25], Myanmar [25], Nepal [21, 26], and Pakistan [25]. The comparison included paratypes, ST, *gyrA* mutations, and phylogenetic relationships among these isolates.

## Results

### Epidemiology of paratyphoid fever

According to NIDSS data from 2001 to 2024, a total of 223 paratyphoid fever cases were reported in Taiwan, with annual case numbers ranging from 0 to 28 and an average incidence of 0.04 per 100,000 population. Among the reported cases, 100 (44.8%) were acquired domestically, while 123 (55.2%) were classified as imported. From 2001 to 2019, 65.2% (120/184) of cases were imported. However, following the COVID-19 pandemic, between 2022 and 2024, 92.1% (35/38) of cases were domestically acquired (Figure 1). Of the 123 imported cases, most originated from Southeast Asia, South Asia, and China, with the top three source countries being Indonesia (40 cases), China (27 cases), and India (22 cases), together accounting for 72.4% of total imported cases. Other countries contributing to imported cases include Cambodia (12), Myanmar (8), Bangladesh (4), Nepal (2), Thailand (2), and several additional countries (5). Imported cases comprised Taiwanese residents who acquired the infection during overseas travel, as well as foreign migrant workers and international travelers who were infected before entering Taiwan.

**Figure 1.**
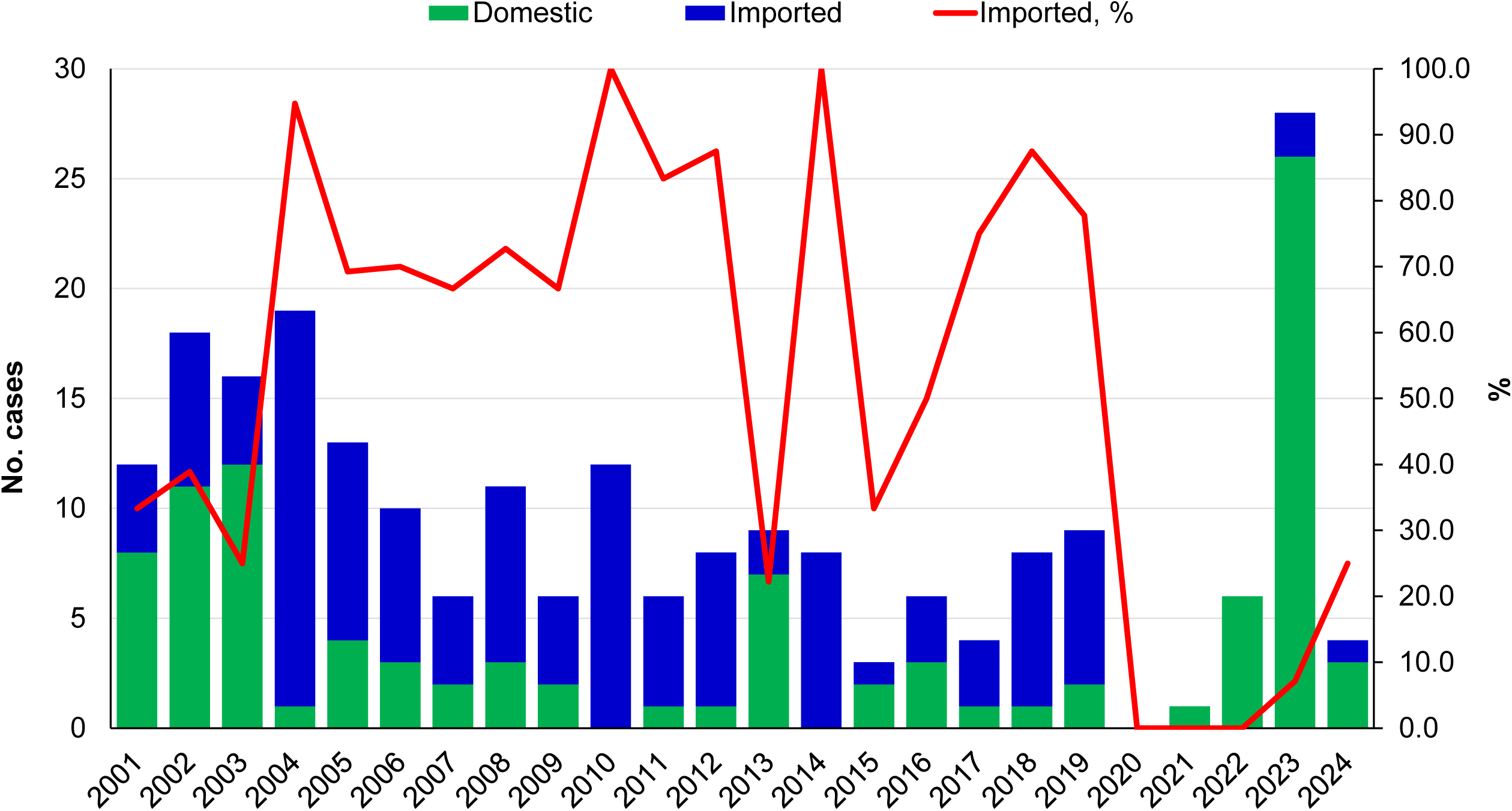
Epidemiological curve of paratyphoid fever cases in Taiwan from 2001 to 2024.

Demographic data were available for 213 of the 223 cases. Among these, females represented 53.5% of total cases, accounting for 50.0% of domestic cases and 56.4% of imported cases. The highest prevalence was observed among young and middle-aged adults, with the peak number of cases in the 25–29 age group, followed by the 30–34 and 35–39 age groups (Figure S1).

### Antimicrobial susceptibility

From 2007 to 2024, a total of 135 paratyphoid fever cases were confirmed in Taiwan, with isolates obtained from 88 cases (65.2%). All isolates were identified as *S.* Paratyphi A. AST revealed that all 88 isolates were susceptible to azithromycin, ampicillin, third-generation cephalosporins (cefotaxime and ceftazidime), meropenem, colistin, gentamicin, sulfamethoxazole, trimethoprim, tetracycline, and tigecycline. However, 76.1% (67/88) of isolates were resistant to nalidixic acid and nonsusceptible to ciprofloxacin, with ciprofloxacin MICs ranging from 0.5 to 1 mg/L among the nalidixic acid-resistant isolates.

### Genomic characteristics and phylogenetic analysis of isolates from Taiwan

WGS was performed on 88 *S.* Paratyphi A isolates (51 imported and 37 domestic) collected between 2007 and 2024. Conventional sequence type analysis identified 26 isolates as ST85 and 62 as ST129. Mutations in *gyrA* were detected in 67 ciprofloxacin-nonsusceptible isolates, with S83F substitution present in 64 isolates and S83Y in 3 isolates, while 21 isolates retained the wild-type *gyrA*. The isolates were classified into 11 paratypes, with the most common being paratype 2.4 (52 isolates), followed by 2.3.1 (8), 2.3.2 (7), and 2.3.3 (7) (Table S1). Among the 37 domestic isolates, 34 were classified as paratype 2.4, which was also identified in 18 of the 19 isolates imported from Indonesia. The remaining three domestic isolates belonged to paratypes 2, 2.3.1, and 2.3.2, respectively. Isolates from Cambodia, China, and Myanmar each corresponded to a single paratype, whereas the 13 isolates from India were distributed across five different paratypes.

A phylogenetic tree based on the cgSNP profiles of 88 isolates revealed distinct clustering patterns corresponding to paratypes and geographic origins (Figure 2). Among the 52 paratype 2.4 isolates, 51, along with one paratype 2 isolate, formed a tight cluster comprising isolates from both Indonesia and Taiwan, suggesting a potential epidemiological link between the two countries in the transmission of paratyphoid fever. Notably, 28 of the 31 domestic isolates collected between 2022 and 2024 exhibited high genetic similarity, sharing paratype 2.4, sequence type ST129, and the *gyrA* S83F mutation. The close genetic relatedness among these isolates indicates a potential outbreak that may have been ongoing since 2022. Of the remaining three domestic isolates detected during the same period, two (R24.0017 and R24.0956) also belonged to paratype 2.4 while harboring wild-type *gyrA*. The third isolate (R23.2261) clustered with paratype 2.3.1 isolates from Cambodia.

**Figure 2.**
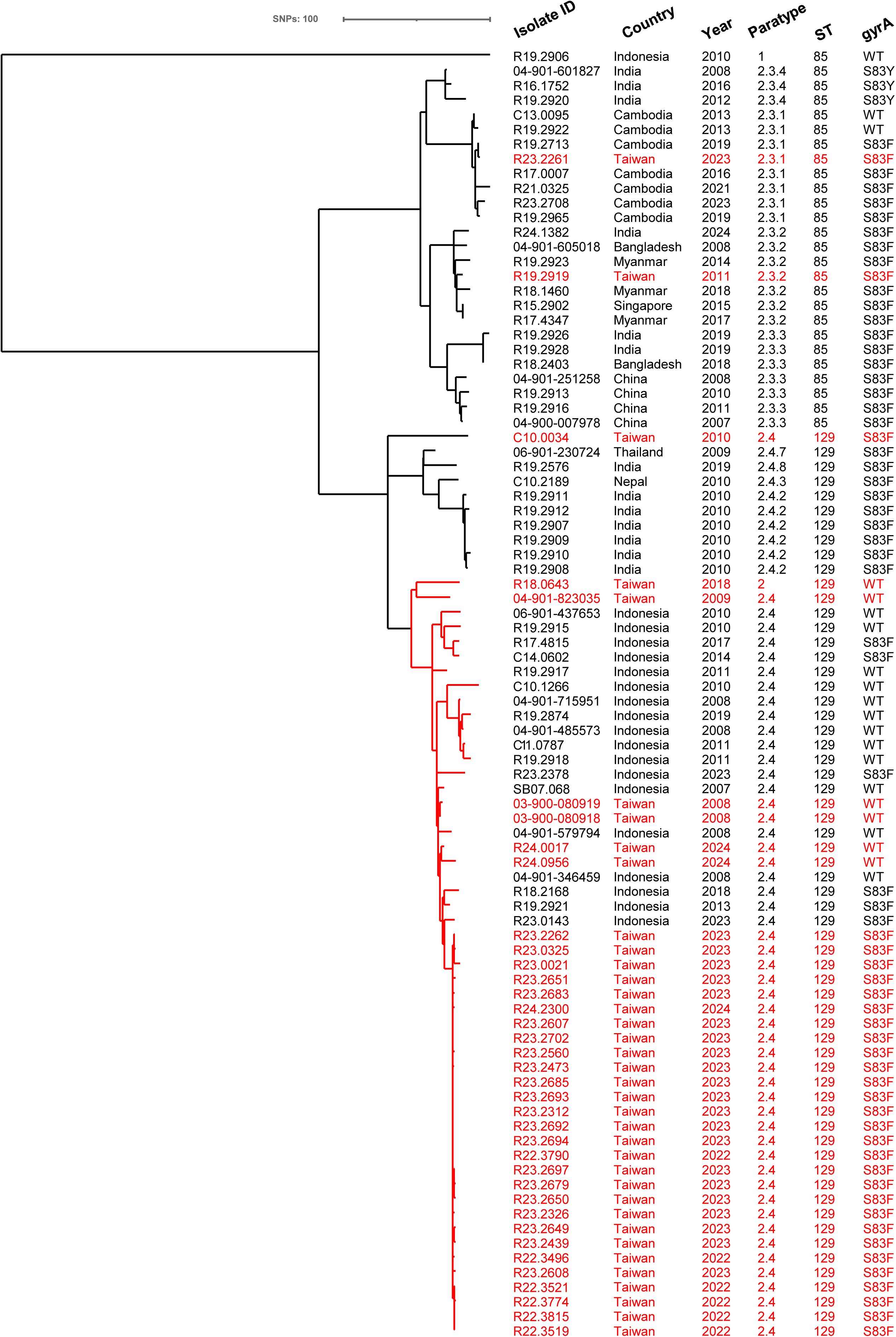
Phylogenetic tree of 88 *S.* Paratyphi A isolates from Taiwan, collected between 2007 and 2024. The tree was constructed based on cgSNP profiles and inferred using the maximum likelihood method. Isolates from domestically acquired cases are highlighted in red. The cluster containing paratypes 2 and 2.4 isolates from Taiwan and Indonesia is outlined in red, excluding one paratype 2.4 isolate that falls outside this cluster. Additional details of the 88 isolates are provided in Table S3.

### Genomic comparison of global isolates

To further explore the origins of the domestic and imported isolates in Taiwan, we compared their genetic relatedness to isolates from Bangladesh, Cambodia, China, India, Indonesia, Myanmar, Nepal, and Pakistan. The genomic analysis revealed that certain countries had predominant paratypes, such as paratype 2.3.1 in Cambodia, 2.3.3 in China, 2.4 in Indonesia, and 2.3.2 in Myanmar (Table S2). In contrast, isolates from Bangladesh, India, Nepal, and Pakistan exhibited greater paratype diversity. The isolates imported from Bangladesh, Cambodia, China, India, Indonesia, Myanmar, and Nepal aligned with the prevalent paratypes in their countries of origin. For example, the 7 isolates imported from Cambodia shared paratype 2.3.1, which was found in 72 of 78 Cambodian isolates. Similarly, the 4 isolates imported from China matched paratype 2.3.3, consistent with 13 other isolates from China. The isolate from Singapore shared paratype 2.3.2, and the one from Thailand matched paratype 2.4.7, commonly found in India. Among the 37 domestic isolates, 34 belonged to paratype 2.4, which is prevalent in Indonesia and also detected in Cambodia, India, Nepal, and Pakistan.

Phylogenetic analysis revealed that isolates within the same paratype generally exhibited a higher genetic relatedness. However, certain paratype 2.3 and 2.4 isolates were genetically distant from the respective predominant clusters (Figure 3). Several paratypes, including 2.3.2, 2.3.3, 2.4, and 2.4.4, formed multiple distinct subclusters that were strongly associated with their geographic origins. For example, imported paratype 2.3.2 isolates from Bangladesh, Myanmar, and Singapore were located in a subcluster, separate from other subclusters containing isolates primarily from India and Nepal. Similarly, a subcluster of paratype 2.3.3 isolates from China, including four from imported cases, was distinct from other subclusters populated by isolates from Bangladesh, India, Nepal, and Pakistan. The paratype 2.4 cluster, which also included one paratype 2 isolate, contained two major subclusters. Subcluster A consisted of isolates from Pakistan and one 2010 isolate (C10.0034) from Taiwan, while subcluster B primarily comprised isolates from Indonesia and Taiwan (Figure S2). Additionally, four paratype 2.4 isolates from Bangladesh and India were genetically distant from both subclusters. All domestic paratype 2.4 isolates identified between 2022 and 2024 showed high genetic similarity to Indonesian isolates. Notably, two 2024 isolates retained the wild-type *gyrA* allele and were more genetically distinct from other isolates carrying the *gyrA* S83F mutation, suggesting differences in their evolutionary trajectories or epidemiological origins.

**Figure 3.**
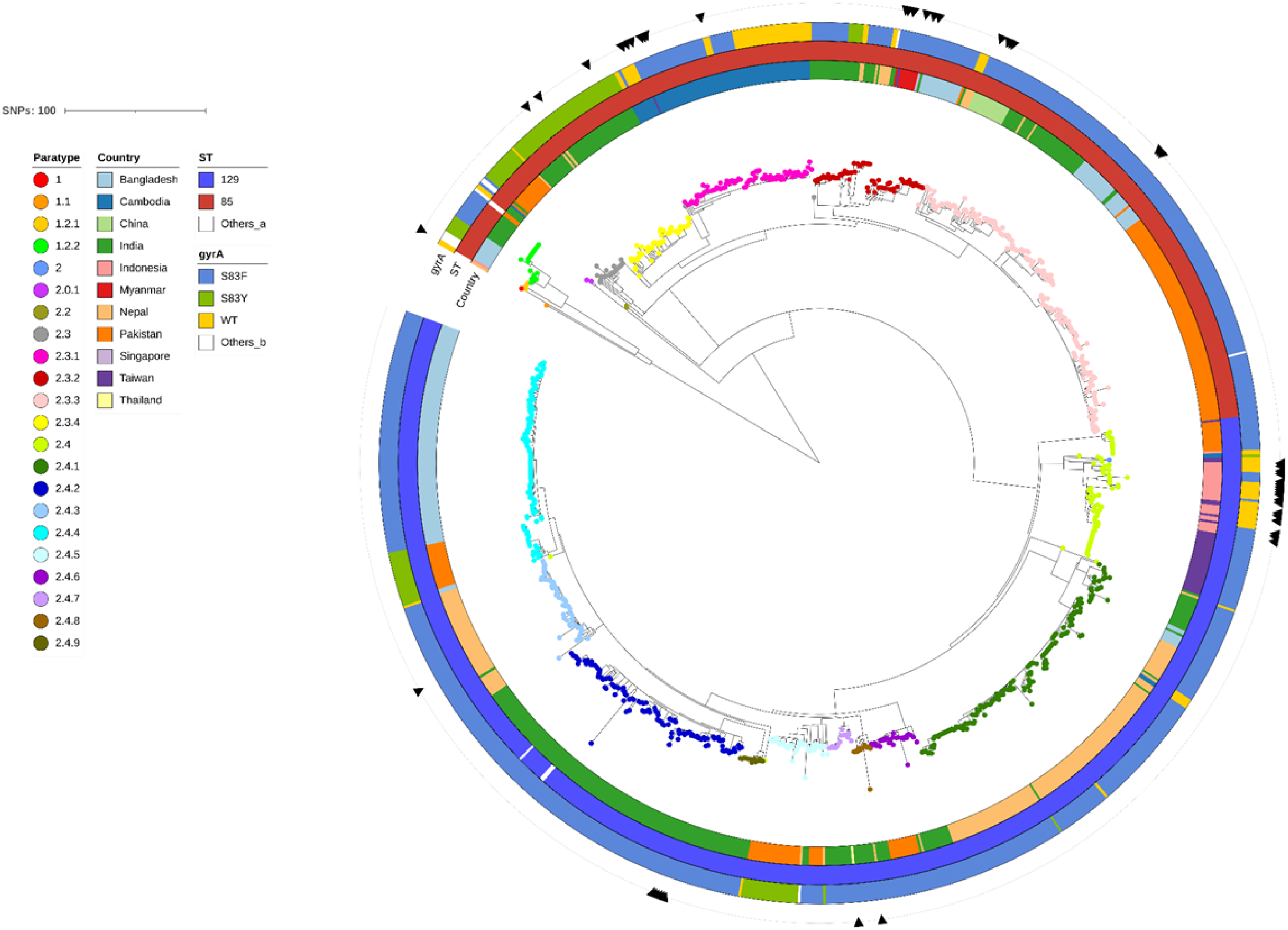
Phylogenetic tree of *S.* Paratyphi A isolates from various countries. The tree was constructed using cgSNP profiles and inferred by maximum likelihood estimation. Isolates collected in Taiwan are marked with arrows in the outermost circle. Additional details of the isolates are provided in Table S4.

## Discussion

This study highlights a notable shift in the epidemiology of paratyphoid fever in Taiwan, marked by a rise in domestically acquired infections since 2022. Before 2022, most cases were imported, primarily from Southeast and South Asia, aligning with the trend observed between 1987 and 1996 [11]. However, from 2022 to 2024, domestic cases outnumbered imported ones, raising concerns about the emergence of local transmission.

Preliminary epidemiologic investigations, although not detailed in this report, revealed that local cases identified since 2022 lacked clear temporal, geographic, or interpersonal connections. Most affected individuals reported recent seafood consumption, suggesting that environmental contamination, possibly involving seafood or water sources, may play a role in transmission. Genomic analysis indicates that most domestic isolates form a tight cluster with Indonesian isolates, suggesting a potential epidemiological link between the two countries in the transmission of paratyphoid fever. Among the 31 domestic isolates collected between 2022 and 2024, 30 clustered with Indonesian strains, and 28 exhibited high genetic similarity, sharing features such as paratype 2.4, ST129, and the *gyrA* S83F mutation. The strong genetic relatedness points to a common source of infection. In the absence of spatiotemporal clustering or direct epidemiological links among cases, the combined findings suggest repeated exposure to a shared food or environmental source rather than ongoing local transmission or a single point source outbreak. A similar pattern was observed during a paratyphoid fever outbreak in Mie Prefecture, Japan, in 1993, where consumption of raw oysters contaminated by *S.* Paratyphi A in harbor waters was identified as the probable source of infection [27]. Notably, the Japanese outbreak exhibited a pattern of geographically dispersed cases without person-to-person transmission, suggesting that environmental exposure played a central role and provided a relevant point of comparison for interpreting the current findings.

Among the 88 *Salmonella* Paratyphi A isolates analyzed in this study, 67 (76.1%) carried mutations in *gyrA*, including 64 with the S83F substitution and 3 with the S83Y substitution. These mutations were associated with resistance to nalidixic acid and reduced susceptibility to ciprofloxacin. This finding is consistent with reports from Bangladesh, Cambodia, and India, where single-point mutations in *gyrA* at codons 83 or 87 are commonly linked to decreased fluoroquinolone susceptibility [6–8]. Although studies from China, India, and Japan have identified additional mutations within the quinolone resistance-determining region as well as plasmid-mediated resistance genes such as *aac(6’)-Ib-cr* [24–26], none were detected in our isolates. All isolates remained susceptible to the other 13 antimicrobials tested, including traditional first-line agents such as ampicillin, chloramphenicol, and trimethoprim-sulfamethoxazole, as well as third-generation cephalosporins, macrolides, aminoglycosides, carbapenems, and others. These results suggest that fluoroquinolone resistance, primarily driven by *gyrA* point mutations, remains the predominant type of antimicrobial resistance among *S.* Paratyphi A isolates collected in Taiwan.

While paratype 2.4 was predominant among both Taiwanese and Indonesian isolates, cgSNP-based phylogenetic analysis revealed considerable genetic divergence within this paratype (Figure S2). For example, paratype 2.4 isolates from Pakistan, India, and Nepal were not only genetically distant from those from Taiwan and Indonesia but also from each other. Although isolates classified under the same paratype generally showed higher genetic similarity compared to those of different paratypes, several exceptions were noted. These findings suggest that the Paratype scheme may not provide sufficient resolution to distinguish between strains with distinct geographic origins. In contrast, cgSNP-based whole-genome comparisons offer a higher resolution, enabling differentiation between strains that belong to the same paratype but are genetically distinct.

## Conclusion

Genomic analysis revealed that *Salmonella* Paratyphi A isolates from recent domestically acquired infections in Taiwan are closely related to strains from Indonesia. Most domestic isolates recovered since 2022 are ciprofloxacin-nonsusceptible, belong to sequence type ST129 and paratype 2.4, and show high genetic similarity, suggesting repeated exposure to a common external source such as contaminated food or environmental reservoirs. These findings highlight the critical role of genomic surveillance in tracing infection sources and facilitating timely public health responses, particularly in food safety and environmental monitoring. The continued susceptibility of these isolates to traditional first-line antibiotics supports their use in current treatment strategies.

## ACKNOWLEDGMENT

This study was supported by the Ministry of Health and Welfare, Taiwan (grant numbers MOHW114-CDC-C-315-124112).

## DATA AVAILABILITY

The whole genome sequences of 88 *S.* Paratyphi A isolates analyzed in this study have been deposited in the National Center for Biotechnology Information under the BioProject accession number PRJNA478278. The specific accession numbers for the isolates are provided in Supplemental Material Table S3.

